# Brain State-dependent Repetitive Transcranial Magnetic Stimulation for Motor Stroke Rehabilitation: A Proof of Concept Randomized Controlled Trial

**DOI:** 10.1101/2024.03.10.24304040

**Authors:** Wala Mahmoud, David Baur, Brigitte Zrenner, Arianna Brancaccio, Paolo Belardinelli, Ander Ramos-Murguialday, Christoph Zrenner, Ulf Ziemann

## Abstract

**Background:** In healthy subjects, repetitive transcranial magnetic stimulation (rTMS) targeting the primary motor cortex (M1) demonstrated plasticity effects contingent on electroencephalography (EEG)-derived excitability states, defined by the phase of the ongoing sensorimotor μ-oscillation. The therapeutic potential of brain state-dependent rTMS in the rehabilitation of upper limb motor impairment post-stroke remains unexplored.

**Objective:** Proof-of-concept trial to assess the efficacy of rTMS, synchronized to the sensorimotor µ-oscillation, in improving motor function and reducing upper-limb spasticity in stroke patients.

**Methods:** We conducted a parallel group, randomized double-blind controlled trial in 30 chronic stroke patients. The experimental intervention group received EEG-triggered rTMS of the ipsilesional M1 (1,200 pulses; 0.33 Hz; 90% of the resting motor threshold (RMT)), while the control group received low-frequency rTMS of the contralesional motor cortex (1,200 pulses; 1 Hz, 115% RMT), i.e., an established treatment protocol. Both groups received 12 rTMS sessions (20 min, 3x per week, 4 weeks) followed by 50 min of physiotherapy. The primary outcome measure was the change in upper-extremity Fugl-Meyer assessment (FMA-UE) scores between baseline, immediately post-treatment and 3 months’ follow-up.

**Results:** Both groups showed significant FMA-UE improvement and spasticity reduction (clinical and objective measures). There were no significant differences between the groups in any of the outcome measures.

**Conclusions:** The application of brain state-dependent rTMS for rehabilitation in chronic stroke patients is feasible. This pilot study demonstrated that the brain oscillation-synchronized rTMS protocol produced beneficial effects on motor disability and spasticity that were comparable to those observed with an established therapeutic rTMS protocol.

## Introduction

In recent years, repetitive transcranial magnetic stimulation (rTMS) has emerged as a safe and non-invasive neuromodulatory intervention for enhancing functional recovery after stroke. One fundamental principle of stroke rehabilitation employing rTMS is to enhance the excitability of the ipsilesional cortex. Two distinct stimulation protocols have been employed for that purpose: 1) High-frequency excitatory rTMS of the ipsilesional hemisphere, with the aim of direct enhancement of the corticospinal output; 2) low-frequency inhibitory rTMS of the contralesional hemisphere to restore excitability balance between the two cortical hemispheres ^1–3^.

There have been some promising findings for the application of low-frequency 1 Hz rTMS to the contralesional hemisphere for functional recovery ^4–6^ and reduction of spasticity ^7^. However, the results of various studies utilizing different rTMS protocols for stroke rehabilitation have not been consistent ^5,8^. The effects, when found, are small and quite variable ^9,10^, indicating that optimal stimulation protocols and parameters are not yet determined. The conventional ’one-size-fits-all’ approach to rTMS therapy has been criticized for neglecting individual patient characteristics. Thus, a shift towards individualized and patient-tailored rTMS therapies is advocated as a solution to reduce heterogeneity and to increase the overall therapeutic efficacy ^11,12^.

Until recently, clinical research involving rTMS has employed an open-loop stimulation approach, disregarding the brain’s instantaneous state at the time of stimulation. The oscillatory activity of the neural networks represents rhythmic fluctuations in excitability ^13–15^, and significantly modulates the network’s response to various inputs ^16–20^. Therefore, taking into account the ongoing oscillatory brain state represents a compelling prospect for optimization of therapeutic brain stimulation.

Several investigations that explored EEG phase-dependent responses to TMS of the human primary motor cortex (M1) revealed that the trough/early rising phase of the endogenous sensorimotor μ-rhythm corresponds to a high-excitability state of corticospinal neurons, as indicated by motor evoked potentials (MEPs) of a larger amplitude compared to the peak of the µ-rhythm ^21–24^. Importantly, these alternating excitability states are decisive for the induction of plasticity: consistent triggering of rTMS during the high-excitability state led to long-term potentiation (LTP)-like effects in corticospinal excitability. In contrast, no changes were observed when stimuli were uncoupled from sensorimotor μ-phase in otherwise identical stimulation protocols ^23,25–27^. In particular, the synchronization of TMS bursts with the trough of the μ-oscillation demonstrated efficacy in LTP-like plasticity induction in healthy subjects when presented in high-gamma frequencies (e.g., 100 Hz) ^28^.

Brain state-dependent TMS has predominantly been explored in healthy subjects. The method relies on autoregressive forward prediction to estimate future instantaneous oscillatory phases ^23,29^, requiring consistent and predictable phase progression over time. In stroke, the disruption of brain networks can undermine the integrity of the μ-oscillation ^30^, which makes reliable phase targeting challenging. Nevertheless, Hussain et al. (2020) ^31^ have recently demonstrated in three chronic stroke patients that accurate targeting of the ipsilesional sensorimotor µ-rhythm is feasible.

Collectively, these findings suggest that real-time information about instantaneous brain states can be utilized to control the efficacy of plasticity in humans, potentially optimizing stimulation protocols to maximize the benefits of therapeutic interventions using rTMS in rehabilitation after stroke. Therefore, the objective of this study was to investigate the feasibility and therapeutic efficacy of µ-phase-synchronized rTMS over M1 in improving motor disability and reducing spasticity of the upper limb in chronic stroke patients. In this proof-of-concept trial, we compared the effects of μ-oscillation-triggered rTMS of the ipsilesional M1 with an established protocol of low-frequency rTMS over the contralesional M1.

## Methods

### Trial design

This single-center parallel-group randomized double-blind controlled proof-of-concept trial compared brain-oscillation-triggered rTMS, synchronized with the trough of the EEG sensorimotor µ-rhythm of the ipsilesional hemisphere (referred to as intervention), with the established protocol of 1 Hz rTMS of the contralesional M1 (referred to as control). Both stimulation protocols were administered immediately prior to upper limb physiotherapy. Blinding procedures were implemented to ensure that participants and study staff, except for the personnel delivering the rTMS treatment, were unaware of group assignment.

The trial was conducted at Tübingen University Hospital-Department of Neurology & Stroke. All patients provided informed consent prior to participating in the trial (ethics committee approval No: 530/2019BO1). The study adhered to the principles of the declaration of Helsinki and was prospectively registered in a publicly accessible clinical trials registry (ClinicalTrials.gov, identifier: NCT05005780).

### Subjects

We recruited patients with upper limb hemiparesis following ischemic or hemorrhagic stroke. The inclusion criteria were as follows: 1) minimum 6 months since stroke onset; 2) age 18– 85 years and able to provide informed consent; 3) presence of ipsilesional MEPs; 4) resting motor threshold (RMT) of the contralesional M1 <80% of maximum stimulator output (MSO); 5) ability to understand and willingness to follow FMA-UE instructions; 6) FMA-UE score ≤60.

Following safety and ethics guidelines for TMS in clinical practice and research ^32^, participants were excluded if they 1) had a seizure disorder history; 2) were taking pro-convulsive medication; 3) were taking muscle-relaxing medication (e.g. baclofen, tolperison, cannabis); 4) had a cardiac pacemaker, implanted medication pump, or an intracranial implant; 5) received a botulinum toxin injection in their affected upper limb <3 months before inclusion; 6) had a wrist joint contracture hindering spasticity measurement.

### Randomization

Patients were randomized using a stratified block randomization method ^33^ with randomly selected block sizes of 4 or 6 patients. Stratification, based on FMA-UE scores, categorized patients into two strata: scores 0-30 and scores 31-60, ensuring a balanced distribution at baseline. The randomized list was generated using MATLAB.

### Study overview

Patients underwent four evaluation sessions. The first session involved eligibility screening, followed by baseline assessment, immediate post-therapy assessment, and a three-month follow-up. The study’s outcome measures were evaluated during all three assessment sessions. Adverse events were systematically monitored through inquiries after each rTMS session, addressing effects or complaints from the current or previous sessions.

Additionally, magnetic resonance images (MRIs) (clinical or research scans, T1 or T2-weighted) were available for a subset of patients (n=24; 11 in the control group and 13 in the intervention group), and used for lesion mapping. Lesions were overlaid for each intervention group (Fig 1C), guiding their classification as subcortical, cortical, or combined, with motor cortex involvement defining cortical lesions (Table S1).

**Fig 1.**
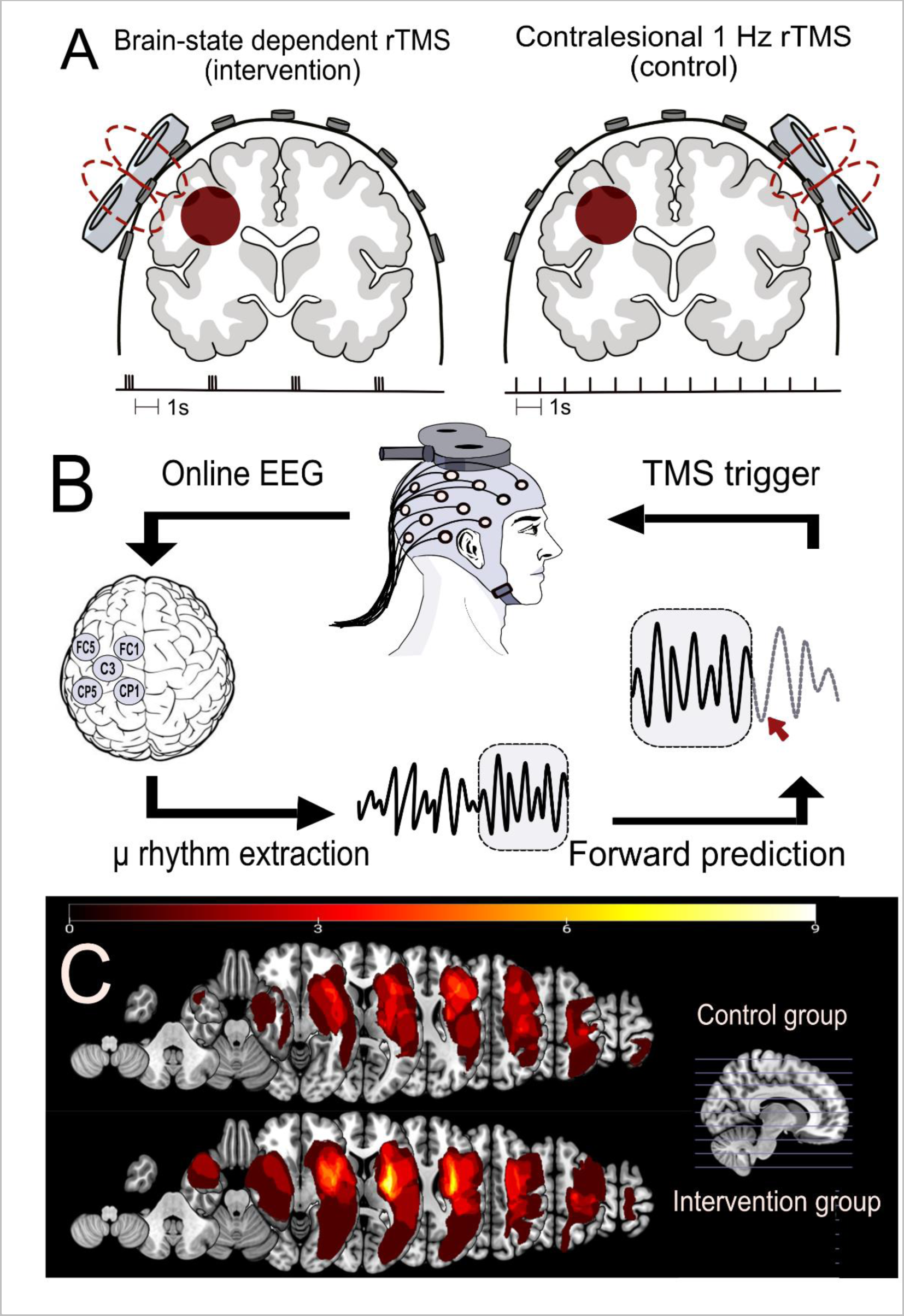
A. The two rTMS protocols used in this study. Left: the brain state-dependent stimulation group (intervention group) received 1,200 TMS pulses to the ipsilesional M1 in the form of triple pulses at 100 Hz and an interstimulus interval of on average 3 s. Right: the control group received 1,200 TMS pulses to the contralesional M1 at 1 Hz frequency. **B.** The brain state-dependent stimulation method: an output copy of the EEG signal was analyzed in real-time in order to determine the timing of and trigger the TMS pulse. The local cortical activity of the sensorimotor cortex μ-rhythm was extracted using a surface Laplacian source derivation montage centered around C3 for a left-hemisphere lesion. A sliding 500 ms window of EEG data was band-pass filtered (6-15 Hz) to isolate the µ-rhythm. An autoregressive model was used to predict the instantaneous phase of the signal. When the band-power exceeded a predetermined threshold and simultaneously matched a pre-specified phase, a TMS pulse was triggered. **C.** Lesion distribution overlays for both study groups, the control group (upper panel, n = 11) and the intervention group (lower panel, n = 13), were generated through MRI analysis. The computation involved manual delineation conducted by a qualified neurologist for each patient to define the Volume of Interest (VOI) within the subject space. Subsequently, the VOI was normalized to Montreal Neurological Institute (MNI) space using the Clinical Toolbox in SPM12. VOIs were mirrored to the left in right hemisphere lesions. Lesion overlap was calculated utilizing the NiiStat toolbox functions for MATLAB.

### TMS stimulation protocols

Both groups received 12 rTMS therapy sessions over 4 weeks (3 sessions per week). During rTMS stimulation sessions, patients were seated on an electronically adjustable reclining chair with both arms relaxed. A conventional TMS stimulator (MagPro R30, MagVenture, Farum, Denmark) was used to deliver biphasic TMS pulses to the motor hotspot, using an air-cooled figure of eight coil (MCF-B65 MagVenture, diameter 75 mm) according to the designated protocol. The coil was oriented such that the second phase of the biphasic pulse induced an electrical field in the brain from lateral-posterior to medial-anterior.

In the 1Hz rTMS group (control group), 1,200 TMS pulses were delivered at a frequency of 1 Hz and an intensity equivalent to 115% RMT to the motor hotspot of the contralesional M1 ^8^. For the brain state-dependent stimulation group (intervention group), phase-specific TMS pulses were triggered by a customized real-time signal-processing system ^23^. Triple pulses at 100 Hz, with an inter-pulse interval of on average 3 s were delivered based on the instantaneous oscillatory phase of the ipsilesional sensorimotor µ-rhythm ^28^. A total of 400 triplets at 90% RMT were triggered using a combined criterion of μ-phase (triggered at negative peak) and power threshold. The power threshold was individually adjusted to result in a stimulation frequency around 0.33 Hz (Fig 1A).

Immediately following each rTMS session, patients engaged in a 50-minute personalized exercise-based physiotherapy training targeting arm and hand function. The training involved practicing meaningful functional tasks tailored to the patients’ goals and motor abilities, covering various aspects like trunk control, strength training, object manipulation, and fine motor training.

### EEG and EMG recordings

NeurOne Tesla biosignal amplifier (Bittium Biosignals Ltd., Finland) was used for acquiring both EEG and EMG signals. Scalp EEG was recorded using a TMS-compatible 64-channel Ag/AgCl sintered ring electrode cap (EasyCap GmbH, Germany) arranged according to the International 10-10 system with a denser electrode array over the sensorimotor cortex ^34^. EEG signal was sampled at a rate of 5 kHz and low pass filtered (1.25 kHz cut-off). EMG signal was sampled at 10 kHz and low pass filtered (1.5 kHz cut-off) using bipolar surface electrodes (Ambu® Neuroline 720, Denmark) placed on the abductor pollicis brevis (APB), first dorsal interosseous (FDI) and abductor digiti minimi (ADM) in a belly-tendon arrangement.

Resting-state EEG data (5 min) was acquired during screening, post-treatment, and follow-up sessions. Before the recording, the skin was carefully prepared by mild skin abrasion (Nuprep Skin Prep Gel, USA). To ensure signal quality, care was taken to maintain electrode impedances below 10 kΩ. Patients were instructed to keep their eyes open and to maintain their neck, jaw, and arm muscles relaxed during the recording.

### Motor hotspot identification and resting motor threshold measurement

The motor hot-spot was determined by identifying the coil position and orientation over the M1 hand representation that elicited the largest and most consistent MEPs in either ABP, FDI or ADM muscles of the affected upper limb (intervention group) or unaffected upper limb (control group). Once identified, the motor hot spot was marked on the EEG cap to ensure consistent coil positioning throughout the stimulation session.

To determine the stimulation intensity, we assessed the RMT in the FDI, APB and ADM muscles. RMT was defined as the minimum stimulus intensity required to elicit MEPs with a peak-to-peak amplitude of at least 50 µV in the target muscle (the muscle with the lowest RMT) in at least 5 out of 10 consecutive TMS pulses delivered to the motor hotspot ^35^.

### Real time stimulation method

EEG-phase synchronization was achieved using a real-time EEG data analysis method described in Zrenner et al. (2018)^23^. Briefly, an algorithm implemented in Simulink Real-Time (Mathworks Ltd, USA, R2017b) was used to analyze an online output copy of the EEG signal in real-time in order to determine the timing of and trigger the TMS pulse. The local cortical activity of the sensorimotor cortex μ-rhythm was extracted using a surface Laplacian source derivation montage centered at electrode C3 for left-sided lesions (referenced to the average of the surrounding electrodes CP1, CP5, FC1, and FC5), and C4 for right-sided lesions (referenced to CP2, CP6, FC2, and FC6). A sliding window (length 500 ms) of data was band-pass filtered (6-15 Hz) to isolate the µ rhythm. This relatively wide range was chosen as individual peak frequencies of the ipsilesional sensorimotor cortex spanned a wider range than the typical alpha band. The instantaneous phase at the time of the TMS trigger decision was estimated by forward-predicting the signal using an autoregressive model. The spectral band-power within the sliding window of data was also estimated in real-time using Fourier Analysis. When the band-power exceeded a predetermined threshold and simultaneously matched a pre-specified phase, a TMS pulse was triggered (Fig 1B). The power threshold was adjusted continuously during the experiment to achieve an average stimulation frequency of 0.33 Hz.

### Outcome measures

The primary outcome measure was defined as the change in motor impairment measured using the standardized FMA-UE ^36^ both immediately post-treatment (Post), and three months post-treatment (Follow-up) in comparison to Baseline.

The Wolf Motor Function Test (WMFT) consisting of 15 timed tasks was employed to evaluate time duration and quality aspects of upper limb function ^37^. Spasticity-related measurements were obtained using the modified Ashworth scale (MAS). The total MAS score was calculated by summing individual scores from 14 movements of multiple upper limb joints. Objective assessment of spasticity utilized a hand-held dynamometer which distinguished stretch-mediated from passive components of the response to muscle stretch (supplementary material 1A). In addition, we measured post-activation depression, a spinal mechanism, which has been found to correlate with spasticity in the upper limb (supplementary material 1B). Patient-centered outcome measures included the visual analogue scale (VAS) for perceived spasticity, the Motor Activity Log (MAL-30) for upper limb use, and the Disability Rating Scale for assessing the impact of spasticity on daily activities.

### Spectral estimation

Spectral estimation was performed on the resting state EEG data obtained during screening sessions using the multi-taper method, applying three distinct tapers (window functions) to baseline-corrected epochs with a length of 1.4 seconds and a 50% overlap. The aperiodic fractal background component of the spectrum was estimated using the irregular-resampling auto-spectral analysis (IRASA) method ^38^ with factors 1.1 to 2.9 in steps of 0.1 and excluding 2.0, and removed from the full spectrum.

The resulting ratio between the power of the periodic component and the aperiodic background noise can be termed signal-to-noise ratio (SNR). SNR determines the maximum achievable precision of phase targeting ^39^. The individual µ-peak frequency was determined from the corrected spectrum in the range between 6 and 15 Hz of both the ipsilesional and contralesional hemispheres, and the respective SNR at that frequency was obtained.

### Real-time EEG phase estimation accuracy

Phase estimation accuracy for each subject was calculated using the resting state EEG dataset. We compared the real-time—causal—phase estimation, where only data preceding the time of interest, i.e., the TMS pulse is available, against a “benchmark” estimation method using the PHASTIMATE toolbox ^39^. The benchmark —non-causal—estimation uses data before and after the time point of interest for instantaneous phase and amplitude determination.

Both (simulated) real-time phase estimate and the non-causal benchmark phase were obtained from TMS-artifact-free resting-state EEG data. For details, please refer to Zrenner et al. (2020) ^40^. Briefly, resting-state EEG data was segmented into overlapping 1.4 s epochs. The phase at the center of each epoch was estimated using both methods. The causal real-time algorithm utilized a 500 ms segment of data preceding the center of the epoch, while the non-causal benchmark method utilized the entire epoch for estimation. Epochs with an alpha oscillation spectral power below the median were discarded from the phase accuracy analysis, to simulate the effect of the real-time amplitude threshold. We then computed the average error between the trial-wise causal and benchmark estimates. Circular standard deviation quantified the variance of trial-wise errors, providing a measure of estimation precision.

### Statistical analysis

Statistical analyses were performed on the intention-to-treat (ITT) population using IBM-SPSS Statistics software version 28.0.1.1 (IBM Corp., Armonk, NY, USA). Baseline comparability between the intervention and control groups included the examination of several variables, namely age, time since stroke, and baseline FMA-UE scores using one-way analysis of variance (ANOVA).

For the primary efficacy analysis, we employed repeated measures ANOVA, considering Session (Baseline, Post, Follow-up) as a within-subject effect, and Group (Intervention, Control) as a between-subject effect. Additionally, was examined the interaction (Session X Group). To assess normality assumptions, we inspected histograms for each outcome measure and calculated skewness and kurtosis. The assumption of sphericity was tested using Mauchly’s test, which indicated no violation of sphericity for any variable, eliminating the need for correction. For ordinal and non-normally distributed variables (such as VAS, Disability Rating Scale), Whitney U nonparametric test was applied. A p-value of <0.05 was considered to indicate statistical significance.

## Results

79 subjects were screened between October 2019 and March 2022. Thirty stroke patients (23 males, 7 females; average age 57.9 ± 8.0 years) participated in this study. All patients had unilateral hemiparesis due to stroke at least six months before participation (average 51 ± 47 months). A flow chart of participants is presented in Fig 2. The characteristics of the study participants are presented in Table S1. Two patients (control group) were lost to 3 months follow-up and one additional patient refused to perform the WMFT during follow-up.

**Fig 2.**
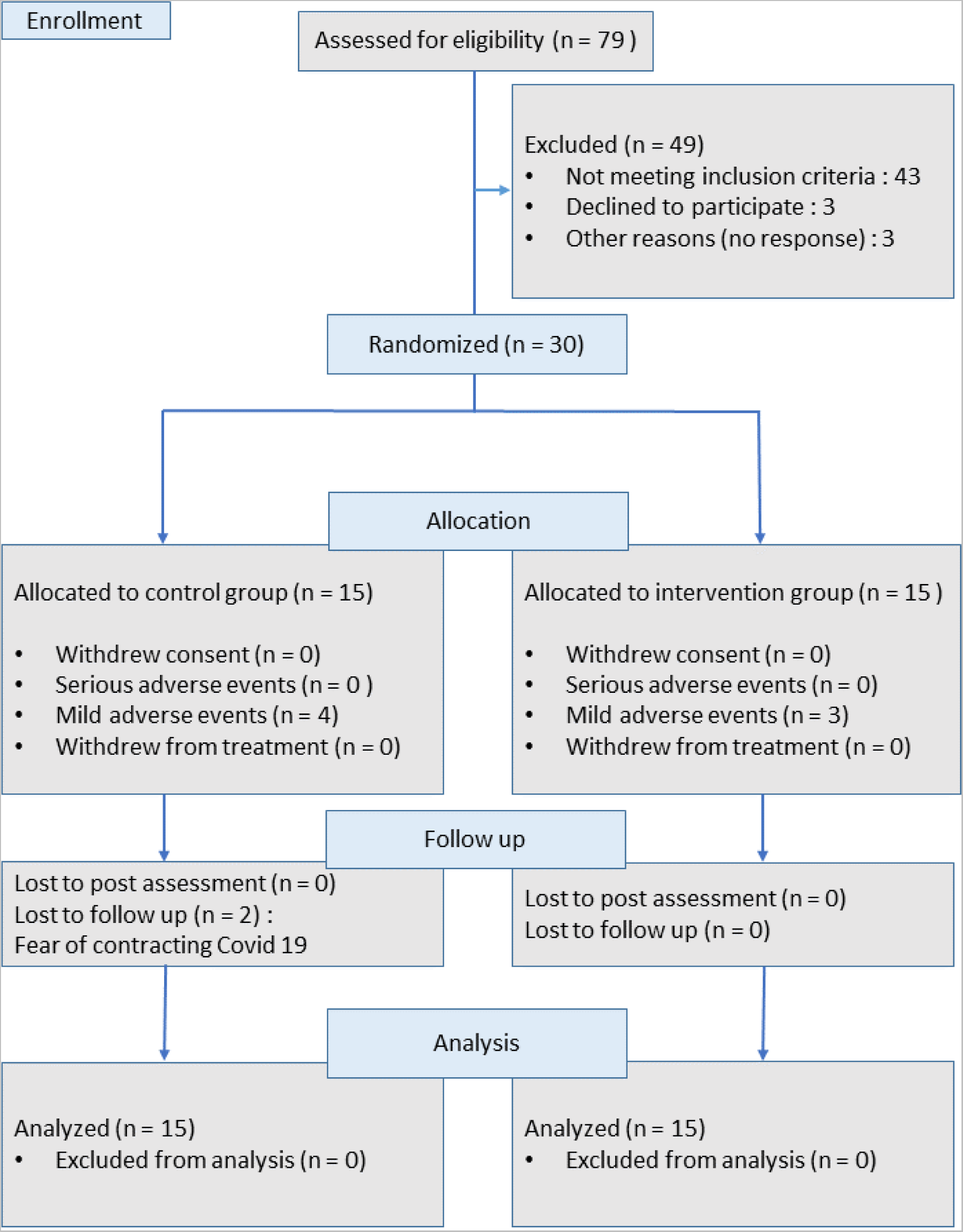
**Flow chart of study participants**

### Adverse events

During the study, adverse events were observed in 7 out of 360 therapy sessions: 4 in the control group and 3 in the intervention group. In the control group, adverse events included tiredness (n=1), headache (n=1), and shoulder/arm pain— presumably, posture, not rTMS-related — (n=2, 1 session terminated). In the intervention group, adverse events consisted of head numbness (n=1), transient double vision (n=1), and tiredness (n=1). Two screening sessions had to be terminated—one due to orthostatic syncope and another due to intense agitation during TMS.

### Spectral analysis and phase estimation accuracy

The mean µ-peak frequency of the ipsilesional sensorimotor cortex was 9.5 ± 2.7 Hz with a mean SNR of 5.4 ± 2.3 dB, compared to 12.5 ± 2.3 Hz and 4.9 ± 2.3 dB in the contralesional sensorimotor cortex. With regards to targeting the µ-oscillation in the ipsilesional hemisphere, the average phase difference between the estimate of the real-time algorithm and the post-hoc phase measure was 1° ± 83.6° (mean ± circular standard deviation) demonstrating an absence of a systematic bias. These results are similar to findings reported in the literature ^31,40^.

### Baseline group comparison

The ANOVA results revealed no significant differences between the two groups at baseline with regards to age (intervention: 57 ± 8 years, control: 60 ± 9 years; p = 0.39), time since stroke (intervention: 47 ± 52 months, control: 54 ± 44 months; p = 0.72), and FMA-UE scores (intervention: 37.6 ± 15, control: 38 ± 15; p = 0.92).

### Effects on clinical outcomes

The repeated measures ANOVA (Table 1) revealed a significant effect of the treatment on FMA-UE score (Session: F _(2,52)_ = 25.7, p < 0.001). No significant effect was observed for Group (F _(1,26)_ = 0.45, p = 0.51) or the Session X Group interaction (F _(2,52)_ = 0.31; p = 0.73). Post hoc analyses indicated significant increases of the FMA-UE score Post vs. Baseline (p < 0.001) and 3-month Follow-up vs. Baseline (p < 0.001), but no difference between Post and Follow-up scores (p=0.48) (Fig. 3).

**Fig 3.**
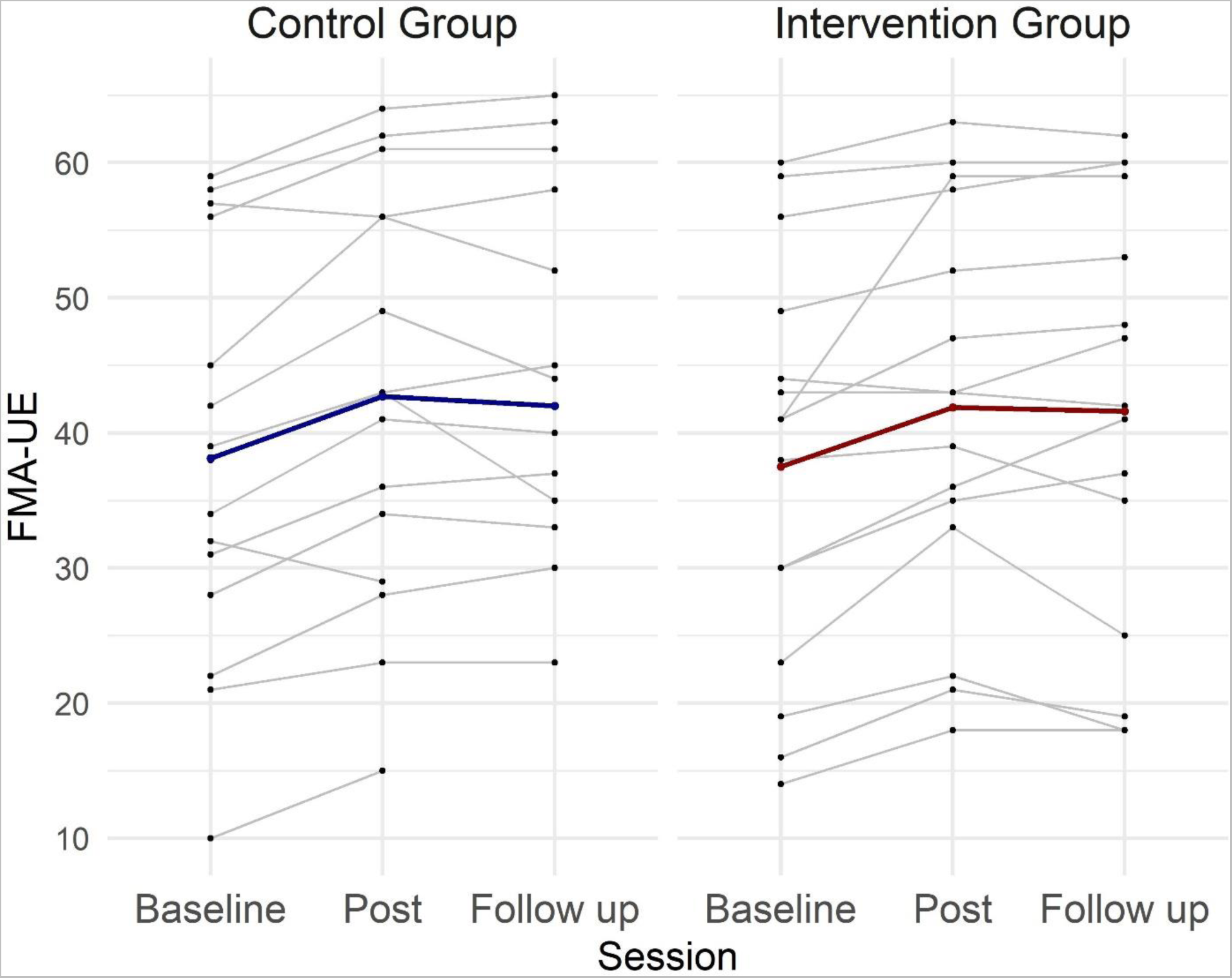
Individual and group FMA-UE scores across sessions for the intervention and control groups. Each thin line represents one patient, while the thick lines represent the mean values for each group across the three evaluation sessions.

**Table 1.** Results of the statistical analysis for clinical outcome measures.

| | **Contralesional 1Hz rTMS**<br>**(Control)**<br>Mean ± standard deviation | | | **Phase-dependent rTMS**<br>**(Intervention)**<br>Mean ± standard deviation | | | | | |
| --- | --- | --- | --- | --- | --- | --- | --- | --- | --- |
| Measure | Pre | Post | Follow-up | Pre | Post | Follow-up | Session | Group | Session X Group |
| FMA-UE / 66 | 40.8*± 3.9 | 45.8 ± 4.0 | 45.1 ± 4.1 | 37.6 ± 3.7 | 41.6 ± 3.7 | 41.6 ± 3.9 | $F_{(2,52)} = 25.7$,<br>$p < 0.001$ | $F_{(1,26)} = 0.45$,<br>$p = 0.51$ | $F_{(2,52)} = 0.31$, $p = 0.73$ |
| WMFT-Time (s) | 652 ± 135 | 518 ± 126 | 564 ± 132 | 731 ± 125 | 659 ± 116 | 681 ± 121 | $F_{(2,50)} = 7.4$, $p = 0.002$ | $F_{(1,26)} = 0.09$, $p = 0.76$ | $F_{(2,50)} = 0.76$, $p = 0.48$ |
| WMFT-function/ 75 | 42.8 ± 5.7 | 46.8 ± 5.7 | 46.7 ± 6 | 37.9 ± 5.2 | 40.2 ± 5.2 | 40.2 ± 5.6 | $F_{(2,48)} = 13.3$,<br>$p < 0.001$ | $F_{(1,24)} = 0.12$, $p = 0.73$ | $F_{(2,48)} = 0.52$, $p = 0.59$ |
| Grip strength (Kg) | 15.3 ± 2.1 | 15.6 ± 2.4 | 15.8 ± 2.0 | 9.0 ± 2.0 | 9.4 ± 2.2 | 8.5 ± 1.9 | $F_{(2,44)} = 0.09$,<br>$p = 0.91$ | $F_{(1,22)} = 5.7$, $p = 0.03$ | $F_{(2,44)} = 0.21$, $p = 0.81$ |
| Stretch reflex torque (mV) | 1.53 ± 0.34 | 1.29 ± 0.28 | 1.58 ± 0.33 | 1.29 ± 0.34 | 0.93 ± 0.28 | 0.92 ± 0.33 | $F_{(1,22)\dagger} = 5.5$, $p = 0.029$ | $F_{(1,22)} = 0.8$, $p = 0.38$ | $F_{(1,22)} = 0.86$, $p = 0.36$ |
| MAS_total / 70 | 9.8 ± 1.3 | 8.2 ± 1.3 | 8.1 ± 1.4 | 8.3 ± 1.2 | 6.6 ± 1.2 | 7.5 ± 1.3 | $F_{(2,50)} = 5.8$, $p = 0.005$ | $F_{(1,25)} = 0.48$, $p = 0.49$ | $F_{(2,50)} = 0.51$, $p = 0.60$ |
| MAL (%) | 23.9 ± 5.6 | 36.9 ± 7 | 31.6 ± 6.6 | 24.9 ± 4.9 | 31.0 ± 6 | 33.8 ± 5.8 | $F_{(2,42)} = 11.6$,<br>$p = < 0.001$ | $F_{(1,21)} = 0.01$, $p = 0.92$ | $F_{(2,42)} = 2.1$, $p = 0.13$ |

Abbreviations: FMA-UE: Fugl-Meyer Assessment-Upper Extremity; WMFT - Wolf Motor Function Test; MAS - Modified Ashworth Scale; MAL-Motor Activity Log.
\* The discrepancy in group mean values between the baseline group comparison analysis and this table is due to the loss of two patients to follow-up. ANOVA systematically excludes individuals with missing data, resulting in means calculated considering only subjects with a complete dataset.
† The ANOVA was run on Session including two levels (Pre, Post) disregarding Follow-up, because data from 10 subjects were lost (5 in each group due to a technical issue and failure to attend the Follow-up session in 2 cases).

The WMFT showed a significant treatment effect (Session) for both the time (F _(2,50_ = 7.4, p = 0.002) and function components (F _(2,48)_ = 13.3, p < 0.001). No Group effect or significant Session X Group interactions were observed. Moreover, there was a significant treatment effect on the reduction of stretch reflex torque measured with the hand-held dynamometer (Session: F _(2,36)_ = 5.5, p= 0.029), without a significant Group effect or Session X Group interaction (Fig. 4).

**Fig 4.**
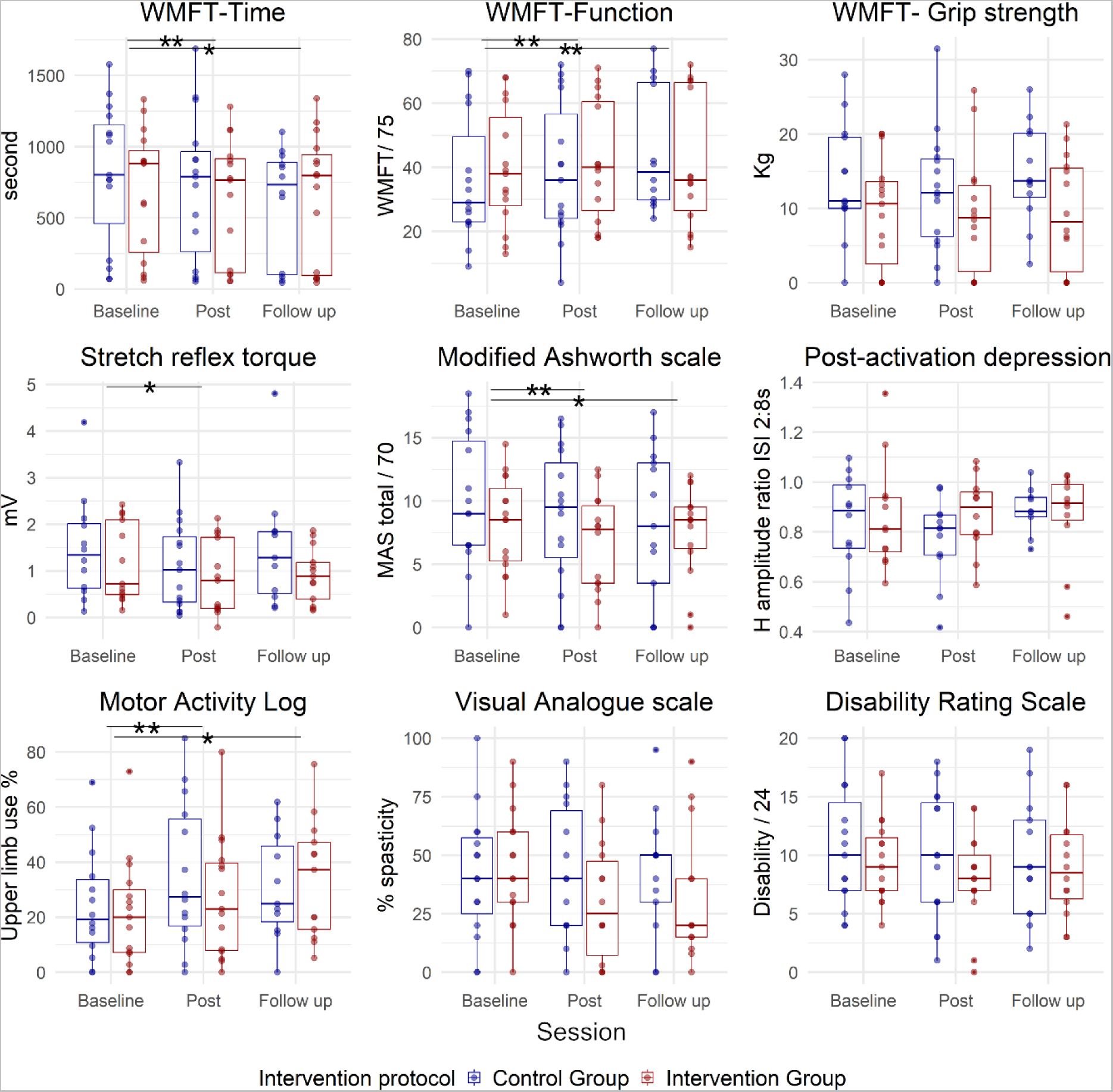
Box plots depicting group data for the two study groups: contralesional 1Hz rTMS (control) group in blue and phase-dependent (intervention) group in red, across three measurement points (pre, post, and follow-up). The small blue and red circles represent individual patient data. Each panel corresponds to one outcome measure. Abbreviations: WMFT - Wolf Motor Function Test; MAS - Modified Ashworth Scale; ISI - Interstimulus Interval. The asterisk represents a significant effect of the treatment (Session) on both groups, with no Session X Group interaction, (*) is significance at p<0.05 level, while (**) represents significance at p<0.001 level.

The MAS (total) exhibited a significant decrease after treatment (Session: F _(2,50)_ = 5.8, p= 0.005), without a significant Group effect or Session X Group interaction. The patient-reported use of the upper extremity in everyday activities (MAL-30) significantly increased after treatment (Session: F _(2,42)_ = 11.6, p< 0.001) without a significant Group effect or Session X Group interaction. No significant effects of the treatment were observed on grip strength, post-activation depression, VAS for perceived spasticity, or Disability Rating Scale (Fig. 4).

## Discussion

Evidence from experiments in healthy subjects indicates that brain state-dependent rTMS may improve the magnitude and consistency of rTMS effects by delivering the stimulation during periods of increased cortical excitability, thus, increasing the induction of LTP-like plasticity ^23,26^. In this trial, we investigated feasibility and efficacy of an rTMS protocol which synchronized the TMS bursts with the trough of the ipsilesional sensorimotor cortex μ-rhythm, i.e., a high-excitability state of the corticospinal tract. We compared this intervention to standard low-frequency rTMS of the contralesional motor cortex.

The stimulation was well-tolerated in both groups. All adverse events were mild, mostly standard TMS side-effects or unspecific in nature. The accuracy of targeting the trough of the ipsilesional sensorimotor cortex μ-rhythm was comparable to previous studies in healthy subjects ^23,27,40^, confirming the feasibility of safe and successful administration of brain state-dependent stimulation to chronic stroke patients in a clinical setting. A more detailed analysis of EEG parameters and changes will be published in a separate manuscript.

The analysis of clinical endpoints demonstrated significant improvements in motor disability and function, alongside a reduction in clinical and objective measures of spasticity in both groups with no significant difference between the groups. Motor improvement was maintained three months’ post-treatment, and was accompanied by an increased use of the affected upper limb in daily activities, as indicated by the MAL questionnaire. This feasibility study, designed with limited statistical power, is not intended for comparing the effectiveness of both stimulation protocols. However, it can serve as a pilot study to inform larger clinical trials. In addition, the absence of a sham-rTMS arm in this study raises the possibility that clinical improvements in both groups may be attributed to the physiotherapy, especially considering the limited physiotherapy typically received by chronic patients. Our study did not aim to establish rTMS superiority over physiotherapy, but rather to compare two rTMS protocols that serve as priming tools for enhancing motor rehabilitation.

Both rTMS protocols implemented in this study are embedded in the interhemispheric inhibition (IHI) model. It supposes that unopposed inhibition from the contralesional to the ipsilesional hemisphere impedes recovery after stroke. The phase-dependent stimulation aimed to directly upregulate the excitability of the ipsilesional cortex using high-frequency (100 Hz) triplets, while the low-frequency (1Hz) rTMS attempted to indirectly upregulate the excitability of the ipsilesional cortex through downregulation of the contralesional hemisphere and thereby rebalancing abnormal interhemispheric inhibition. The integrity of the ipsilesional corticospinal tracts has been proposed as a biomarker for potential benefit from therapies that aim to upregulate ipsilesional excitability ^41^. Despite the different working mechanisms of the two stimulation protocols used in this study, both aimed to enhance the excitability of the ipsilesional hemisphere. In line with this, only patients with positive ipsilesional MEPs were included, thereby increasing both the homogeneity of the study sample and the potential for improvement.

Selection of contralesional low-frequency stimulation as the control intervention was based on the cumulative evidence supporting its efficacy. However, recent literature questions its mechanism and suitability for all stroke patients ^42^. Indeed, the IHI imbalance model itself is currently being debated and re-evaluated in light of recent evidence ^43–47^, suggesting that IHI per se should not be a target for rTMS interventions ^46^. Contemporary models of interhemispheric communication present a more complex picture of the interaction between the two hemispheres following stroke, contingent on factors such as structural reserve ^48^ and integrity of the callosal and frontal connections ^49^. It is crucial to emphasize, however, that the ongoing debate around the IHI model does not invalidate previously reported beneficial effects (for an overview, see: Kim et al. (2020), and Starosta et al. (2022)) ^50,51^. Rather, it suggests that these effects likely arise from different, yet unknown mechanisms ^46^.

Two key principles are proposed to enhance the effectiveness of therapeutic brain stimulation: precision and personalization ^52^. This proof-of-concept study showed that the brain state-dependent stimulation protocol produced outcomes comparable to an established conventional open-loop stimulation protocol. This makes synchronization of brain stimulation to individual brain oscillations a promising possibility for personalizing therapeutic rTMS interventions. It is necessary to acknowledge, however, that this trial is preliminary. Comprehensive investigations are necessary to explore the full potential of this technology. Currently, a confirmatory multicenter randomized controlled trial is ongoing in Germany to test the therapeutic efficacy in subacute motor stroke patients (ClinicalTrials.gov, identifier: NCT05600374) ^53^.

A primary strategy to improve the effectiveness of this approach is through patient selection and stratification. The selection of an ideal rTMS protocol should consider factors like stroke type, location, and the stage of stroke recovery. In addition, the future of therapeutic brain stimulation should encompass enhanced subject stratification through in-depth analysis of brain connectivity and structural integrity. In the context of optimizing phase-dependent stimulation protocols specifically, several considerations arise. Firstly, it may be necessary to choose patients with a sufficiently high µ-rhythm power and SNR for optimal phase targeting ^39^. Secondly, given heterogeneity in lesion location and size among the stroke population, EEG channel selection may be essential. Thirdly, parameters for brain state-dependent stimulation could be entirely automated using personalized classifiers ^54,55^.

### Effects on spasticity

When evaluating the impact of the rTMS therapy on spasticity, our investigation revealed a significant reduction in both MAS-total and the MAS-wrist extension. These results align with previous studies ^56–59^. This study represents one of the first attempts to objectively assess the impact of rTMS on spasticity. Using recently developed technology, we found a significant reduction in the stretch reflex torque, which represents the neurogenic component of resistance to joint stretch. No change was recorded in the passive stiffness components. These findings are similar to what we observed in a previous study ^60^ which exclusively involved patients receiving 1 Hz contralesional rTMS.

Here, we found that high-frequency rTMS of the ipsilesional M1 (brain state-dependent) also reduced spasticity. The finding that both stimulation protocols were similarly able to improve motor function and to reduce spasticity may indicate that reduction of spasticity could be related to motor function improvement, a finding that we recorded previously ^60^. This improvement, which may be reflected in an increased excitability of the ipsilesional corticospinal tract can trigger changes at the level of the spinal cord and possibly the motor neurons itself, which lead to a reduction in the stretch reflex-mediated torque. We also examined the possibility of the involvement of the post activation depression as a putative mechanism underlying the measured reduction in spasticity. This was not confirmed from this data, in line with our previous study, which examined this effect only by applying the 1Hz rTMS protocol to contralesional M1 ^60^.

## Conclusions

This study demonstrates that brain state-dependent stimulation can be successfully applied in chronic stroke patients with clinical effects that are similar to current TMS standard therapy. EEG-TMS therefore constitutes an exciting option for personalization and optimization of motor neurorehabilitation supported by non-invasive brain stimulation. Future studies should further explore its clinical benefits and optimize its application.

## Data Availability

All data produced in the present study are available upon reasonable request to the authors

## Acknowledgments

W.M was funded by the German Academic Exchange Program (DAAD) throughout the period of the project. D.B was supported by an intramural Junior Clinician Scientist grant while B.Z. and C.Z were funded by an EXIST Forschungstransfer grant from the German Federal Ministry for Economic Affairs and Energy. We thank the TMS outpatient clinic team who organized and delivered the stimulation, especially Dragana Galevska and Julia Greilich.

## References

1. Fisicaro F, Lanza G, Grasso AA, et al. Repetitive transcranial magnetic stimulation in stroke rehabilitation: review of the current evidence and pitfalls. Ther Adv Neurol Disord. 2019;12:1756286419878317–1756286419878317. doi:10.1177/1756286419878317

2. Graef P, Dadalt MLR, da Silva Rodrigués DAM, Stein C, de Souza Pagnussat A. Transcranial magnetic stimulation combined with upper-limb training for improving function after stroke: a systematic review and meta-analysis. J Neurol Sci. 2016;369:149–158.

3. Le Q, Qu Y, Tao Y, Zhu S. Effects of Repetitive Transcranial Magnetic Stimulation on Hand Function Recovery and Excitability of the Motor Cortex After Stroke: A Meta-Analysis. Am J Phys Med Rehabil. 2014;93(5). https://journals.lww.com/ajpmr/fulltext/2014/05000/effects_of_repetitive_transcranial_magnet ic.8.aspx

4. Avenanti A, Coccia M, Ladavas E, Provinciali L, Ceravolo MG. Low-frequency rTMS promotes use-dependent motor plasticity in chronic stroke: a randomized trial. Neurology. 2012;78(4):256–264.

5. Lefaucheur JP, Aleman A, Baeken C, et al. Evidence-based guidelines on the therapeutic use of repetitive transcranial magnetic stimulation (rTMS): An update (2014–2018). Clin Neurophysiol. 2020;131(2):474–528. doi:10.1016/j.clinph.2019.11.002

6. Luk KY, Ouyang HX, Pang MYC. Low-Frequency rTMS over Contralesional M1 Increases Ipsilesional Cortical Excitability and Motor Function with Decreased Interhemispheric Asymmetry in Subacute Stroke: A Randomized Controlled Study. Neural Plast. 2022;2022:3815357. doi:10.1155/2022/3815357

7. Wang X, Ge L, Hu H, Yan L, Li L. Effects of Non-Invasive Brain Stimulation on Post-Stroke Spasticity: A Systematic Review and Meta-Analysis of Randomized Controlled Trials. Brain Sci. 2022;12(7):836. doi:10.3390/brainsci12070836

8. Harvey RL, Edwards D, Dunning K, et al. Randomized sham-controlled trial of navigated repetitive transcranial magnetic stimulation for motor recovery in stroke. Stroke. Published online 2018.

9. López-Alonso V, Cheeran B, Río-Rodríguez D, Fernández-del-Olmo M. Inter-individual variability in response to non-invasive brain stimulation paradigms. Brain Stimul. 2014;7(3):372–380.

10. Smith M-C, Stinear CM. Transcranial magnetic stimulation (TMS) in stroke: Ready for clinical practice? J Clin Neurosci. 2016;31:10–14. 10.1016/j.jocn.2016.01.034

11. Koch PJ, Hummel FC. Toward precision medicine: tailoring interventional strategies based on noninvasive brain stimulation for motor recovery after stroke. Curr Opin Neurol. 2017;30(4):388–397. doi:10.1097/WCO.0000000000000462

12. Leuchter AF, Corlier J. A precision medicine approach to repetitive Transcranial Magnetic Stimulation (rTMS). Brain Stimul. 2018;11(3):463–464. doi:10.1016/j.brs.2018.02.003

13. Arieli A, Sterkin A, Grinvald A, Aertsen AD. Dynamics of ongoing activity: explanation of the large variability in evoked cortical responses. Science (80-). 1996;273(5283):1868-1871.

14. Bergmann TO, Mölle M, Schmidt MA, et al. EEG-guided transcranial magnetic stimulation reveals rapid shifts in motor cortical excitability during the human sleep slow oscillation. J Neurosci.2012;32(1):243–253.

15. Huerta PT, Lisman JE. Bidirectional synaptic plasticity induced by a single burst during cholinergic theta oscillation in CA1 in vitro. Neuron. 1995;15(5):1053–1063.

16. Karabanov A, Thielscher A, Siebner HR. Transcranial brain stimulation: closing the loop between brain and stimulation. Curr Opin Neurol. 2016;29(4):397–404. doi:10.1097/WCO.0000000000000342

17. Keil J, Timm J, SanMiguel I, Schulz H, Obleser J, Schönwiesner M. Cortical brain states and corticospinal synchronization influence TMS-evoked motor potentials. J Neurophysiol. 2014;111(3):513–519.

18. Li LM, Violante IR, Leech R, et al. Brain state and polarity dependent modulation of brain networks by transcranial direct current stimulation. Hum Brain Mapp. 2019;40(3):904–915.

19. Thut G, Bergmann TO, Fröhlich F, et al. Guiding transcranial brain stimulation by EEG/MEG to interact with ongoing brain activity and associated functions: a position paper. Clin Neurophysiol. 2017;128(5):843–857.

20. Zrenner C, Belardinelli P, Müller-Dahlhaus F, Ziemann U. Closed-Loop Neuroscience and Non-Invasive Brain Stimulation: A Tale of Two Loops. Front Cell Neurosci. 2016;10:92. doi:10.3389/fncel.2016.00092

21. Schaworonkow N, Caldana Gordon P, Belardinelli P, Ziemann U, Bergmann TO, Zrenner C. μ-Rhythm Extracted With Personalized EEG Filters Correlates With Corticospinal Excitability in Real-Time Phase-Triggered EEG-TMS. Front Neurosci. 2018;12:954. doi:10.3389/fnins.2018.00954

22. Wischnewski M, Haigh ZJ, Shirinpour S, Alekseichuk I, Opitz A. The phase of sensorimotor mu and beta oscillations has the opposite effect on corticospinal excitability. Brain Stimul. 2022;15(5):1093–1100. 10.1016/j.brs.2022.08.005

23. Zrenner C, Desideri D, Belardinelli P, Ziemann U. Real-time EEG-defined excitability states determine efficacy of TMS-induced plasticity in human motor cortex. Brain Stimul. 2018;11(2):374–389.

24. Zrenner C, Kozák G, Schaworonkow N, et al. Corticospinal excitability is highest at the early rising phase of sensorimotor µ-rhythm. Neuroimage. 2023;266:119805. doi:10.1016/j.neuroimage.2022.119805

25. Baur D, Galevska D, Hussain S, Cohen LG, Ziemann U, Zrenner C. Induction of LTD-like corticospinal plasticity by low-frequency rTMS depends on pre-stimulus phase of sensorimotor μ-rhythm. Brain Stimul. 2020;13(6):1580–1587.

26. Hussain SJ, Vollmer MK, Stimely J, et al. Phase-dependent offline enhancement of human motor memory. Brain Stimul. 2021;14(4):873–883.

27. Schaworonkow N, Triesch J, Ziemann U, Zrenner C. EEG-triggered TMS reveals stronger brain state-dependent modulation of motor evoked potentials at weaker stimulation intensities. Brain Stimul. 2019;12(1):110–118.

28. Baur D, Ermolova M, Souza VH, Zrenner C, Ziemann U. Phase-amplitude coupling in high-gamma frequency range induces LTP-like plasticity in human motor cortex: EEG-TMS evidence. *Brain Stimul Basic*, Transl Clin Res Neuromodulation. 2022;15(6):1508–1510.

29. Madsen KH, Karabanov AN, Krohne LG, Safeldt MG, Tomasevic L, Siebner HR. No trace of phase: Corticomotor excitability is not tuned by phase of pericentral mu-rhythm. Brain Stimul. 2019;12(5):1261–1270.

30. Hughes SW, Crunelli V. Thalamic Mechanisms of EEG Alpha Rhythms and Their Pathological Implications. Neurosci. 2005;11(4):357–372. doi:10.1177/1073858405277450

31. Hussain SJ, Hayward W, Fourcand F, et al. Phase-dependent transcranial magnetic stimulation of the lesioned hemisphere is accurate after stroke. *Brain Stimul Basic*, Transl Clin Res Neuromodulation. 2020;13(5):1354–1357.

32. Rossi S, Antal A, Bestmann S, et al. Safety and recommendations for TMS use in healthy subjects and patient populations, with updates on training, ethical and regulatory issues: Expert Guidelines. Clin Neurophysiol. 2021;132(1):269–306. 10.1016/j.clinph.2020.10.003

33. Efird J. Blocked randomization with randomly selected block sizes. Int J Environ Res Public Health. 2011;8(1):15–20.

34. Seeck M, Koessler L, Bast T, et al. The standardized EEG electrode array of the IFCN. Clin Neurophysiol. 2017;128(10):2070–2077.

35. Groppa S, Oliviero A, Eisen A, et al. A practical guide to diagnostic transcranial magnetic stimulation: report of an IFCN committee. Clin Neurophysiol. 2012;123(5):858–882.

36. Sanford J, Moreland J, Swanson LR, Stratford PW, Gowland C. Reliability of the Fugl-Meyer Assessment for Testing Motor Performance in Patients Following Stroke. Phys Ther. 1993;73(7):447–454. doi:10.1093/ptj/73.7.447

37. Wolf SL, Catlin PA, Ellis M, Archer AL, Morgan B, Piacentino A. Assessing Wolf motor function test as outcome measure for research in patients after stroke. Stroke. 2001;32(7):1635–1639.

38. Wen H, Liu Z. Separating Fractal and Oscillatory Components in the Power Spectrum of Neurophysiological Signal. Brain Topogr. 2016;29(1):13–26. doi:10.1007/s10548-015-0448-0

39. Zrenner C, Galevska D, Nieminen JO, Baur D, Stefanou MI, Ziemann U. The shaky ground truth of real-time phase estimation. Neuroimage. 2020;214:116761. doi:10.1016/j.neuroimage.2020.116761

40. Zrenner B, Zrenner C, Gordon PC, et al. Brain oscillation-synchronized stimulation of the left dorsolateral prefrontal cortex in depression using real-time EEG-triggered TMS. Brain Stimul. 2020;13(1):197–205. doi:10.1016/j.brs.2019.10.007

41. Boyd LA, Hayward KS, Ward NS, et al. Biomarkers of stroke recovery: Consensus-based core recommendations from the Stroke Recovery and Rehabilitation Roundtable. Int J Stroke. 2017;12(5):480–493. doi:10.1177/1747493017714176

42. Safdar A, Smith M-C, Byblow WD, Stinear CM. Applications of Repetitive Transcranial Magnetic Stimulation to Improve Upper Limb Motor Performance After Stroke: A Systematic Review. Neurorehabil Neural Repair. 2023;37(11-12):837–849. doi:10.1177/15459683231209722

43. Gerges ANH, Hordacre B, Pietro F Di, Moseley GL, Berryman C. Do Adults with Stroke have Altered Interhemispheric Inhibition? A Systematic Review with Meta-Analysis. J Stroke Cerebrovasc Dis. 2022;31(7):106494. 10.1016/j.jstrokecerebrovasdis.2022.106494

44. McDonnell MN, Stinear CM. TMS measures of motor cortex function after stroke: a meta-analysis. Brain Stimul. 2017;10(4):721–734.

45. Stinear CM, Petoe MA, Byblow WD. Primary motor cortex excitability during recovery after stroke: implications for neuromodulation. Brain Stimul. 2015;8(6):1183–1190.

46. Xu J, Branscheidt M, Schambra H, et al. Rethinking interhemispheric imbalance as a target for stroke neurorehabilitation. Ann Neurol. 2019;85(4):502–513.

47. Carson RG. Inter-hemispheric inhibition sculpts the output of neural circuits by co-opting the two cerebral hemispheres. J Physiol. 2020;598(21):4781–4802. doi:10.1113/JP279793

48. Di Pino G, Pellegrino G, Assenza G, et al. Modulation of brain plasticity in stroke: a novel model for neurorehabilitation. Nat Rev Neurol. 2014;10(10):597–608.

49. Brancaccio A, Tabarelli D, Belardinelli P. A new framework to interpret individual inter-hemispheric compensatory communication after stroke. J Pers Med. 2022;12(1):59.

50. Kim W-J, Rosselin C, Amatya B, Hafezi P, Khan F. Repetitive transcranial magnetic stimulation for management of post-stroke impairments: An overview of systematic reviews. J Rehabil Med. 2020;52(2):1–10.

51. Starosta M, Cichoń N, Saluk-Bijak J, Miller E. Benefits from repetitive transcranial magnetic stimulation in post-stroke rehabilitation. J Clin Med. 2022;11(8):2149.

52. Wessel MJ, Egger P, Hummel FC. Predictive models for response to non-invasive brain stimulation in stroke: A critical review of opportunities and pitfalls. Brain Stimul. 2021;14(6):1456–1466. 10.1016/j.brs.2021.09.006

53. Lieb A, Zrenner B, Zrenner C, et al. Brain-oscillation-synchronized stimulation to enhance motor recovery in early subacute stroke: a randomized controlled double-blind three-arm parallel-group exploratory trial comparing personalized, non-personalized and sham repetitive transcranial magnetic stimulation (Acronym: BOSS-STROKE). BMC Neurol. 2023;23(1):204. doi:10.1186/s12883-023-03235-1

54. Hussain SJ, Quentin R. Decoding personalized motor cortical excitability states from human electroencephalography. Sci Rep. 2022;12(1):6323.

55. Metsomaa J, Belardinelli P, Ermolova M, Ziemann U, Zrenner C. Causal decoding of individual cortical excitability states. Neuroimage. 2021;245:118652. 10.1016/j.neuroimage.2021.118652

56. Chervyakov A V, Poydasheva AG, Lyukmanov RH, et al. Effects of navigated repetitive transcranial magnetic stimulation after stroke. J Clin Neurophysiol. 2018;35(2):166–172.

57. Galvão SCB, Dos Santos RBC, Dos Santos PB, Cabral ME, Monte-Silva K. Efficacy of coupling repetitive transcranial magnetic stimulation and physical therapy to reduce upper-limb spasticity in patients with stroke: a randomized controlled trial. Arch Phys Med Rehabil. 2014;95(2):222–229.

58. Gottlieb A, Boltzmann M, Schmidt SB, et al. Treatment of upper limb spasticity with inhibitory repetitive transcranial magnetic stimulation: A randomized placebo-controlled trial. NeuroRehabilitation. 2021;49(3):425–434.

59. Kuzu Ö, Adiguzel E, Kesikburun S, Yaşar E, Yılmaz B. The effect of sham controlled continuous theta burst stimulation and low frequency repetitive transcranial magnetic stimulation on upper extremity spasticity and functional recovery in chronic ischemic stroke patients. J Stroke Cerebrovasc Dis. 2021;30(7):105795.

60. Mahmoud W, Hultborn H, Zuluaga J, et al. Testing spasticity mechanisms in chronic stroke before and after intervention with contralesional motor cortex 1 Hz rTMS and physiotherapy. J Neuroeng Rehabil. 2023;20(1):150. doi:10.1186/s12984-023-01275-9

